# Accuracy of AI-assisted diagnostic tools for *Schistosoma haematobium*: A systematic review and meta-analysis

**DOI:** 10.1101/2025.10.31.25339255

**Authors:** Sisay Desale, Getaneh Alemu, Tadesse Hailu

**Author notes:** Corresponding author: Contact address of corresponding author, Name: Sisay Desale, P.O. BOX: 1145.

## Abstract

**Background:** Urogenital schistosomiasis caused by *Schistosoma haematobium* remains endemic in sub-Saharan Africa. Diagnosis traditionally relies on urine microscopy to detect parasite eggs; however, its sensitivity declines in low-intensity infections. Artificial intelligence (AI)-assisted image analysis offers a promising approach to automate egg detection and enhance diagnostic accuracy, but its performance compared with standard microscopy is not well established.

**Methods:** We conducted a systematic review following the PRISMA guidelines and checklist. Studies evaluating AI-assisted detection of *S. haematobium* compared with microscopy and/or molecular reference standards, published up to August 2025, were identified through searches in PubMed/MEDLINE, HINARI, Epistemonikos, Science Direct, Google Scholar and grey literature sources. Eligible studies were selected based on pre-defined inclusion and exclusion criteria. The quality of included studies was assessed using the QUADAS-2 tool. Heterogeneity among studies was evaluated using the Cochrane Q test and I² statistic. Data were analyzed using STATA version 14.1 and Review Manager version 5.4.1.

**Results:** A total of ten studies comprising 18 datasets and 7,318 participants met the eligibility criteria. The pooled sensitivity and specificity of AI-assisted diagnostic tools compared with standard microscopy were 0.88 (95% CI: 0.84–0.92) and 0.89 (95% CI: 0.85–0.93), respectively. The pooled diagnostic odds ratio (DOR) was 64 (95% CI: 38–106), and the area under the summary receiver operating characteristic curve (AUC) was 0.95 (95% CI: 0.92–0.96). The positive likelihood ratio (PLR) was 28.86 (95% CI: 12.01– 67.00), while the negative likelihood ratio (NLR) was 0.03 (95% CI: 0.01–0.10). Subgroup analysis revealed higher diagnostic accuracy of AI-assisted tools in community surveys (sensitivity = 0.93, specificity = 0.90) and superior pooled performance for AiDx platforms (sensitivity = 0.93, specificity = 0.91) compared with SchistoScope (sensitivity = 0.86, specificity = 0.86). Overall, substantial heterogeneity was observed across studies (I² = 99%).

**Conclusion:** AI-assisted diagnostic tools demonstrate high accuracy for detecting *S. haematobium* and could serve as valuable complements to existing diagnostic approaches. However, further field validation, standardization and integration into routine diagnostic workflows are needed to ensure their reliability and scalability in real-world settings.

## Background

Schistosomiasis is a parasitic disease caused by trematodes of the genus *Schistosoma* and remains endemic in many tropical and subtropical regions, particularly in sub-Saharan Africa and parts of the Middle East [1, 2]. Over 250 million people are infected with schistosomiasis worldwide [3], predominantly in impoverished tropical regions, and most cases occur in sub-Saharan Africa. Urogenital schistosomiasis caused by *S. haematobium* is endemic throughout Africa and parts of the Middle East [2], reflecting the distribution of *Bulinus* snail hosts. In Ethiopia alone, about 5.0 million people are infected and 37.5 million are at risk [3], underscoring the high regional burden.

The life cycle of *Schistosoma haematobium* involves freshwater snails (*Bulinus* spp.) as intermediate hosts. Miracidia hatch from eggs in human urine and infect snails, where they develop into cercariae. These cercariae are released into water and penetrate human skin during water contact [4]. Within the human host, the worms mature and settle in the venous plexuses of the bladder and ureters. Females lay eggs that become lodged in the bladder wall, provoking a granulomatous immune response that leads to inflammation, ulceration, fibrosis, and calcification of the urinary tract [2, 5]. Consequently, patients often develop hematuria and other urinary tract pathologies. Chronic infection can cause obstructive uropathy, hydronephrosis, and increases the risk of squamous cell carcinoma of the bladder. In women, egg deposition in the genital tract causes female genital schistosomiasis (FGS), characterized by mucosal lesions, contact bleeding, vaginal discharge, and infertility [2, 5–7].

Praziquantel remains the only effective anti-schistosome drug, typically administered in preventive chemotherapy programs targeting school-aged children and high-risk populations [8]. However, the drug does not prevent reinfection or affect immature worms, highlighting the need for integrated approaches, including health education, improved sanitation, and snail control [9].

The WHO’s 2021–2030 NTD roadmap explicitly aims to eliminate schistosomiasis as a public health problem by 2030. Meeting this goal requires highly sensitive diagnostics, especially as prevalence falls. The WHO target product profile for schistosomiasis diagnostics emphasizes the need to reliably detect low-prevalent (3-10%) infections [2]. The gold standard method, urine filtration microscopy, is simple but has low sensitivity in light or post-treatment infections [10]. Reagent-strip tests for hematuria provide a rapid and inexpensive screening option but cannot distinguish *S. haematobium* from other causes of urinary tract bleeding [11]. Molecular techniques such as polymerase chain reaction (PCR) and loop-mediated isothermal amplification (LAMP) offer high sensitivity, detecting as few as 5 parasites/μL, but require laboratory facilities and skilled personnel [12, 13]. Antigen detection assays like the up-converting phosphor lateral-flow (UCP-LF) CAA test demonstrate excellent sensitivity (around 97%) in low-transmission settings but are not yet field-deployable in many endemic areas [14, 15].

As prevalence and intensity decline, the sensitivity of conventional parasitological methods diminishes, leading to underestimation of infection rates [13]. Thus, there is a growing need for novel diagnostic approaches that combine high sensitivity and specificity with affordability and field adaptability.

Recent advances in artificial intelligence (AI) and deep learning have introduced new opportunities for diagnostic automation in parasitology. Convolutional neural networks (CNNs) can be trained to detect parasite eggs from digital microscopy images with high accuracy [16]. Smartphone-based platforms such as SchistoScope use AI to identify *S. haematobium* eggs directly from filtered urine images [17], while automated bench-top scanners (AiDx Assist) utilize deep learning models to quantify egg counts rapidly [18]. AI has also been applied in reading dipstick color changes for hematuria and proteinuria detection with over 97% accuracy [19].

Despite these advances, AI-based diagnostic studies for *S. haematobium* remain few and reported performance metrics vary widely. For instance, sensitivities range from 83% to 96% and specificities from 77% to 99%, depending on image acquisition, AI model, and reference standard used [16, 20]. These inconsistencies require systematic evaluation to generate pooled estimates and guide implementation feasibility. Hence, the objective of this systematic review and meta-analysis was to systematically evaluate the diagnostic accuracy of artificial intelligence assisted tools for detecting *Schistosoma haematobium* infection.

## Methods

### Search strategy and study selection

This systematic review and meta-analysis adhered to PRISMA guidelines [21], and was registered in PROSPERO (2025: CRD420251140737). We searched in PubMed/MEDLINE, HINARI, Epistemonikos, ScienceDirect, Google Scholar, and Google for studies published up to August 2025. Search terms included combinations of “Schistosoma haematobium” OR schistosomiasis) AND (“artificial intelligence” OR “machine learning” OR “deep learning” OR “AI”) AND (diagnosis OR “diagnostic accuracy” OR detect*. Gray literature and the reference lists of included studies were also reviewed to identify additional relevant articles. The search strategy was designed based on the PIRD (Population, AI-assisted diagnostic tools, Reference standard, Diagnosis of interest) framework. Literature searches were conducted for publications from databases from 5-31 August 2025.

Two reviewers (SD and TH) independently screened titles and abstracts to identify potentially eligible studies. The full text of selected articles was then assessed against the inclusion criteria. Any disagreements were resolved through discussion and consensus.

### Study selection

We included original studies evaluating an AI-assisted tool for detecting *S. haematobium* eggs in human urine samples, using microscopy or molecular diagnostics as the reference standard. Both cross-sectional and cohort study designs were eligible. Only peer-reviewed articles published in English were considered. We excluded review articles, editorials, letters, case reports, animal studies, and conference abstracts.

### Data extraction and quality assessment

From each included study, two reviewers independently extracted data using a standardized excel form. We collected information on first authors, year of publication, study setting and population, sample size, details of the AI-assisted diagnostic tools (device or algorithm), reference standard method, and diagnostic accuracy outcomes (numbers of true positives, false positives, true negatives, and false negatives). Discrepancies in data extraction were resolved by discussion. The methodological quality and risk of bias of each study were assessed using the QUADAS-2 (Quality Assessment of Diagnostic Accuracy Studies, version 2) tool, which evaluates bias in patient selection, AI-assisted diagnostic tools, reference standard, and flow and timing.

### Statistical analysis

Pooled accuracy of AI-assisted diagnostic tools for *Schistosoma haematobium* was analyzed against reference tests such as urine filtration microscopy or PCR using STATA version 14.1. The number of participants with true positive (TP), false positive (FP), false negative (FN), and true negative (TN) AI-assisted test results were used to calculate the sensitivity and specificity for each study as well as the overall summary estimates. Positive likelihood ratio (LR^+^), negative likelihood ratio (LR^−^), and diagnostic odds ratio (DOR) were also calculated. Subgroup analyses were performed based on reference standards, endemicity level, and AI algorithm type. To assess the ability of AI-assisted tools to discriminate participants with *S. haematobium* infection from non-infected individuals, a summary receiver operating characteristic (SROC) curve was generated, and the area under the curve (AUC) was interpreted as excellent (0.9–1.0), good (0.8–<0.9), fair (0.7–<0.8), poor (0.6–<0.7), or failed (0.5–<0.6). Between-study heterogeneity was evaluated using Cochrane’s Q test, and the I² statistic, with significant heterogeneity defined as I² > 50% and Q-test P < 0.10. Methodological quality of the included studies was assessed using the QUADAS-2 tool.

## Results

A total of 320 records were initially identified. After removing 76 duplicates, 244 titles and abstracts were screened, leading to the exclusion of 223 records. Twenty-one full-text articles were then assessed for eligibility, of which 11 were excluded (six were not AI-assisted diagnostic accuracy studies and five did not provide sufficient diagnostic data) (Figure 1). A total of 10 studies, comprising 18 datasets and involving 7,318 participants, were included in this review (Table 1). Sample sizes ranged from 68 participants [20], to 869 in a large community-based study [22], with a median of approximately 398 participants. The studies were all conducted in sub-Saharan Africa, reflecting the endemic distribution of *Schistosoma haematobium*. Geographically, four studies were conducted in Nigeria [18, 22–24], three in Côte d’Ivoire [17, 20, 25], and three in Gabon [18]. All included studies employed cross-sectional design. The majority (4 studies) focused on school-aged children [17, 18, 25], while others were community-based studies that included participants aged five years and above [22, 23] or mixed populations including preschool children, school-aged children, and adults [24]. Seven studies were conducted in moderate-to-high endemic settings such as Gabon and Nigeria [18, 22, 24], and three were conducted in lower prevalence communities, in Côte d’Ivoire [17, 20]. This variation allows the applicability of AI-assisted diagnostics across different transmission intensities.

**Figure 1:**
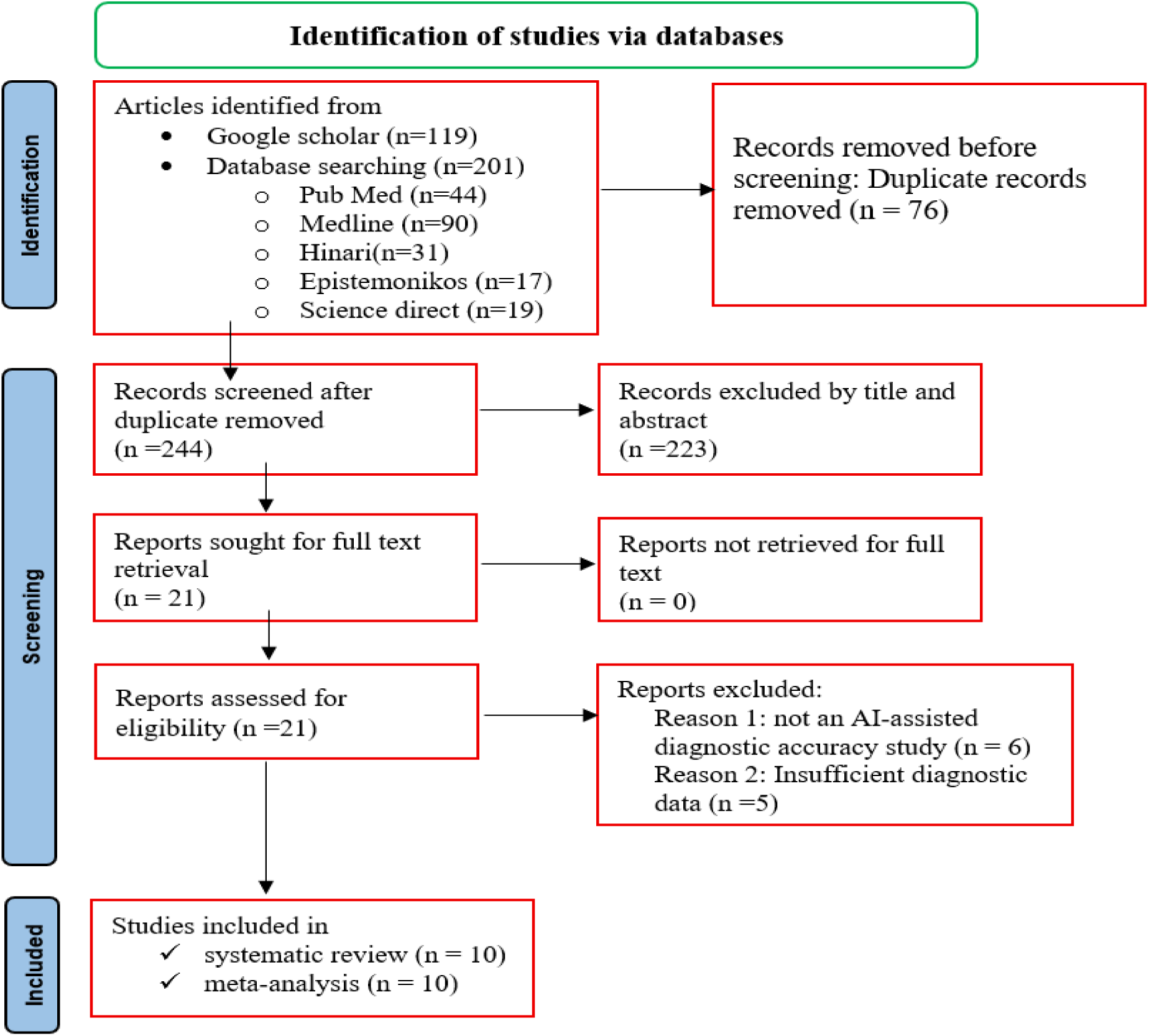
Flow chart showing selection process of eligible studies

**Table 1:**
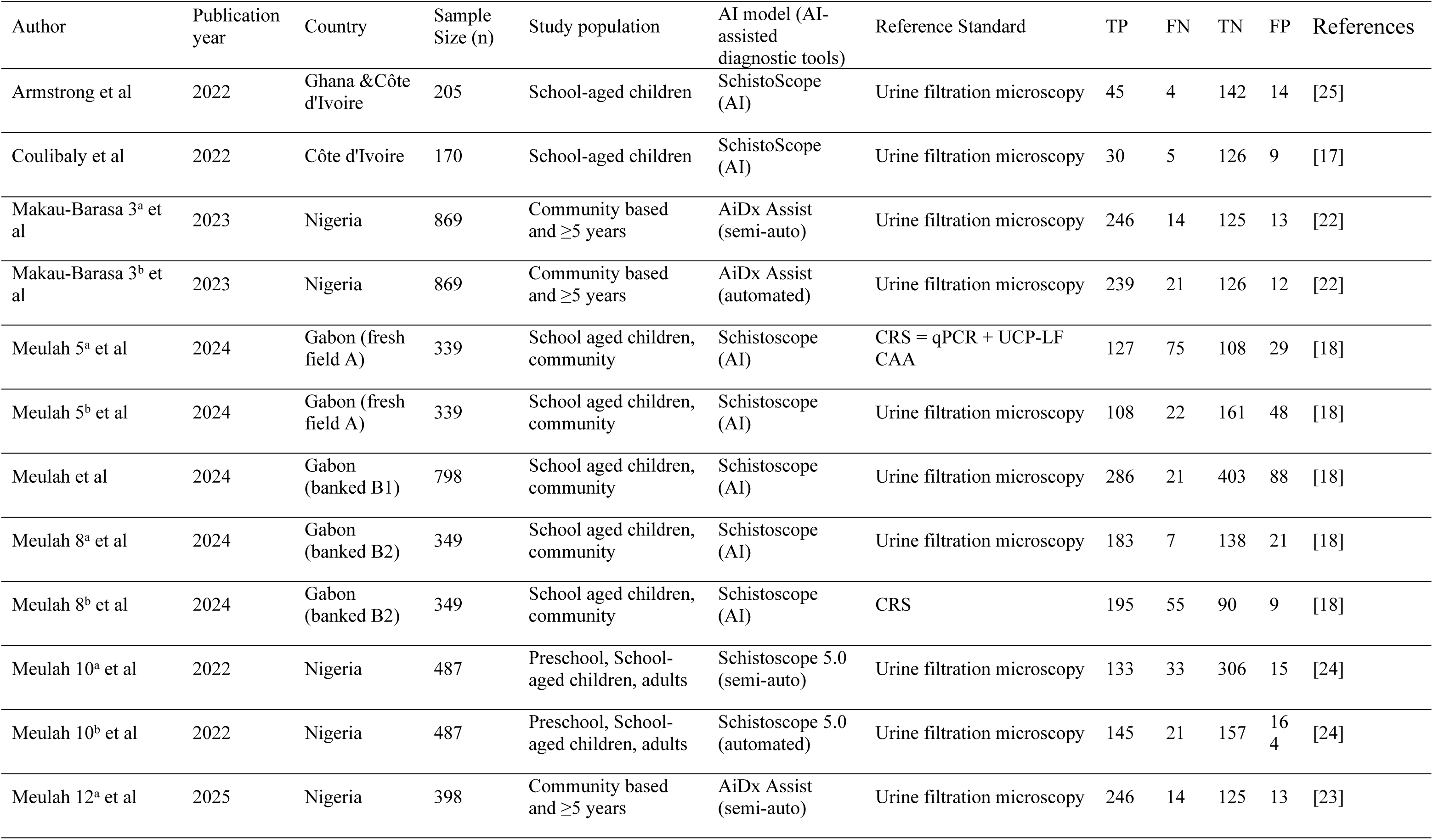
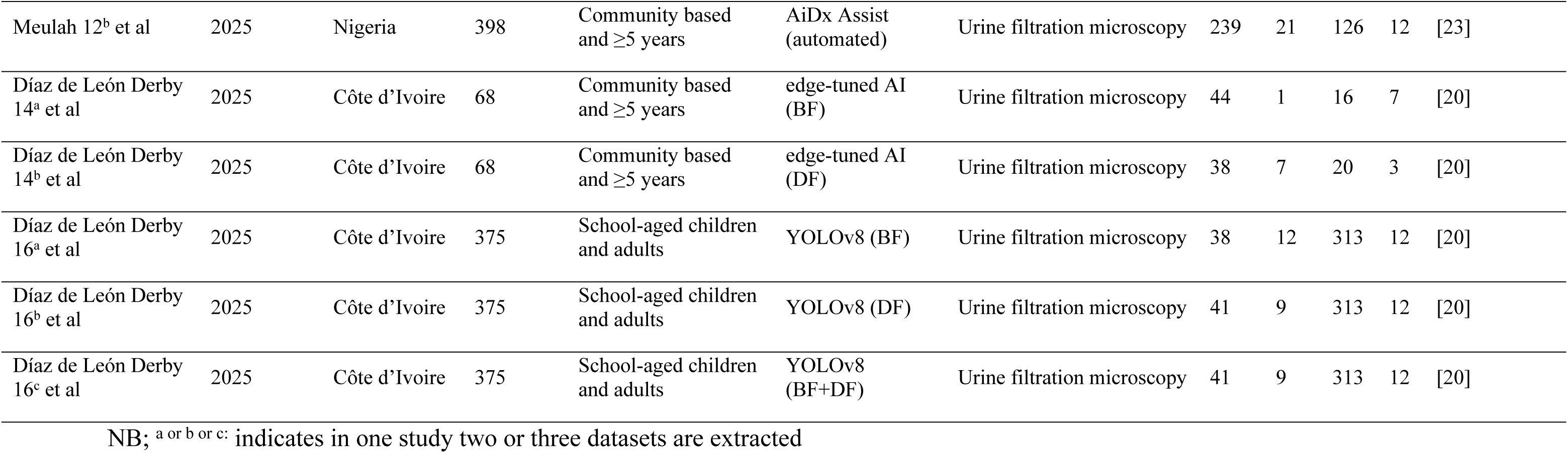
Characteristics of included studies.

Reference standards also varied. Most datasets (n = 8) used conventional urine filtration microscopy as the comparator [17, 18, 20, 22–25]. Two datasets from Gabon additionally employed composite reference standards incorporating qPCR and UCP-LF CAA antigen tests [18]. Urine was consistently the diagnostic specimen across all studies. Some studies processed freshly collected field samples, while others analyzed banked urine samples stored under controlled conditions [18].

Several AI-assisted platforms were assessed across the studies. These included the smartphone-based SchistoScope [17, 18, 25], the AiDx Assist, a semi- and fully automated diagnostic microscope [22, 23], the Schistoscope 5.0, a field-deployable AI-enhanced microscope [24], and more advanced deep learning approaches such as YOLOv8 and edge-tuned AI [20].

### Data quality assessment

The QUADAS-2 evaluation revealed that most studies demonstrated a low risk of bias across the domains of AI-assisted diagnostic tools, reference standard, and flow and timing. However, the patient selection domain showed some limitations, with two studies at high risk of bias and four rated as unclear, mainly due to non-random sampling or insufficient reporting of recruitment methods. Applicability concerns were low across all domains, although a small number of studies raised uncertainties in relation to the AI-assisted diagnostic tools (n=2) and reference standard (n = 2). Overall, the evidence base is moderately robust, but pooled estimates should be interpreted with some caution due to potential bias arising from patient selection in a few studies (Figure 2).

**Figure 2:**
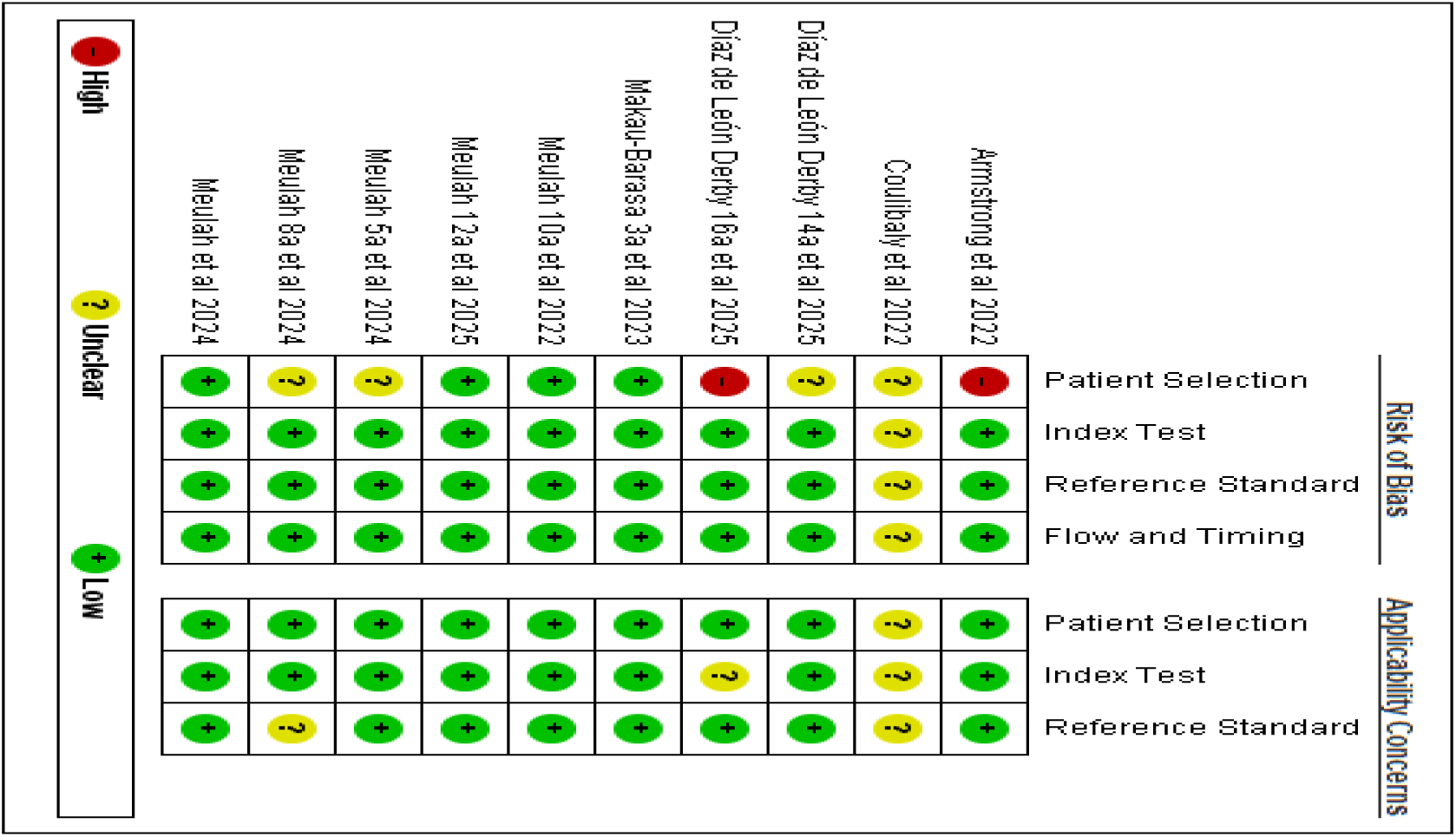
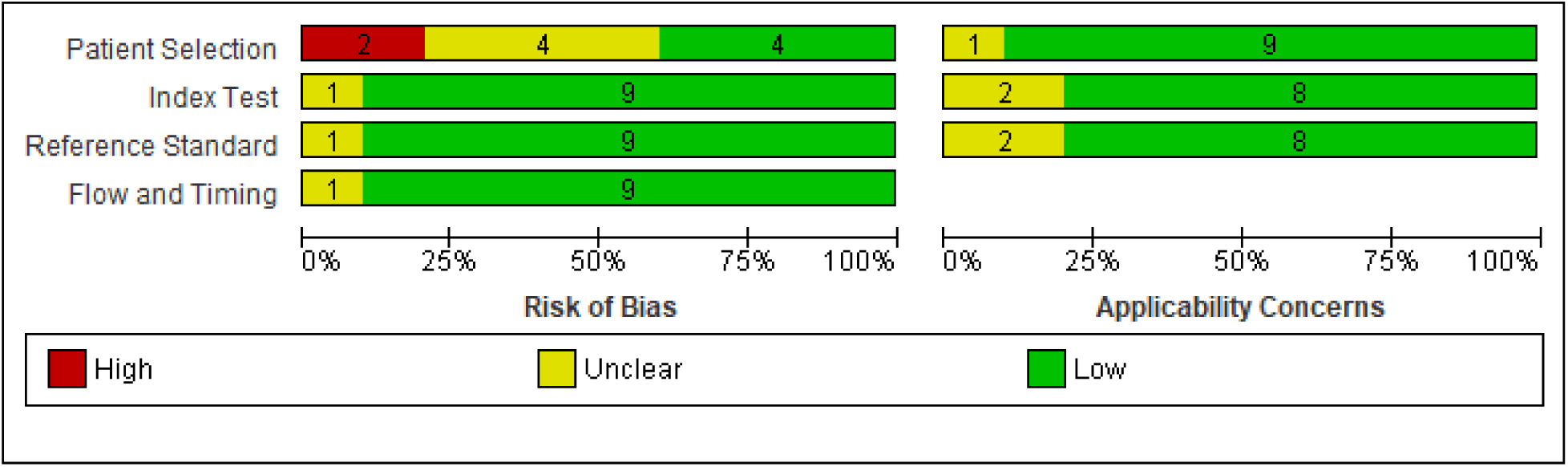
Risk of bias graph of studies included in the meta-analysis analysis.

### Diagnostic accuracy of AI-assisted diagnostic tools

The meta-analysis, which combined all AI-assisted diagnostic tools (including SchistoScope, AiDx Assist, Schistoscope 5.0, and YOLO-based models), demonstrated that the AI-assisted diagnostic tools have high overall diagnostic accuracy. The pooled sensitivity was 0.88 (95%CI: 0.84-0.92) and the pooled specificity was 0.89 (95%CI: 0.85-0.93) (Figure 3). The pooled positive diagnostic likelihood ratio (DLR^+^) of AI-assisted tools for *Schistosoma haematobium* was 28.86 (95%CI: 12.01-67.00), while the pooled negative likelihood ratio (DLR^−^) was 0.03 (95%CI: 0.01-0.10), indicating strong diagnostic performance (Figure 4). The diagnostic odds ratio (DOR) was 64 (95%CI: 38-106), reflecting strong overall discriminatory power. The area under the ROC curve (AUROC) was 0.95 (95%CI: 0.92-0.96), confirming excellent overall performance, distinguishing between infected and non-infected individuals (Figure 5). However, substantial heterogeneity was observed across studies (I² = 99%), particularly in specificity (MED-SPE = 0.69, 95%CI: 0.64-0.76). Therefore, we applied a bivariate random-effects meta-analysis model. Assuming a pre-test probability of 20%, the positive likelihood ratio (LR^+^ = 8) increased the post-test probability of *S. haematobium* infection to 67%, whereas the negative likelihood ratio (LR^−^ = 0.13) reduced it to 3%, indicating reliable discrimination between infected and uninfected individuals (Figure 6).

**Figure 3:**
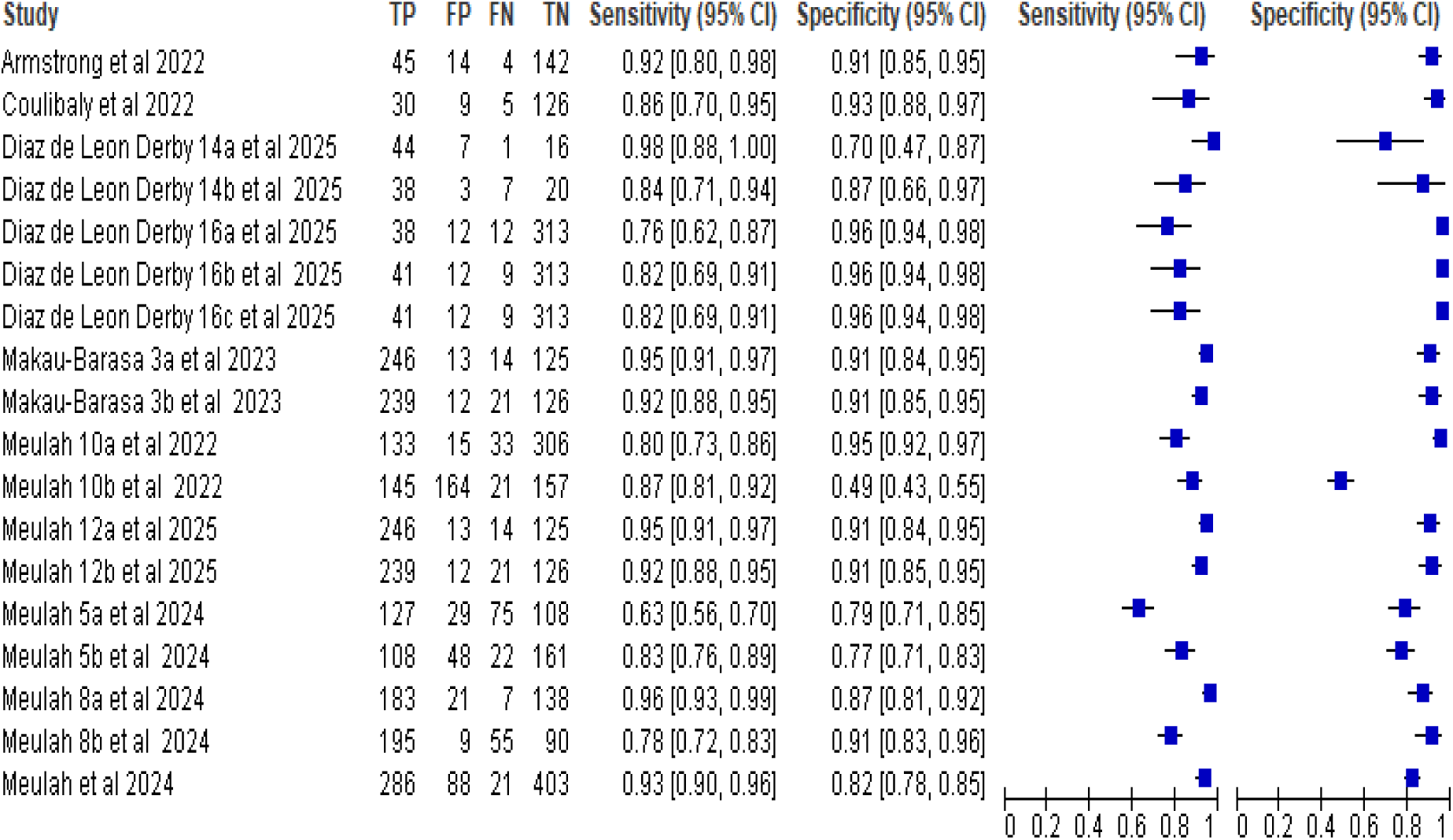
Forest plot illustrates the sensitivity and specificity of AI-assisted diagnostic tools in the discrimination of positive and negative patients.

**Figure 4:**
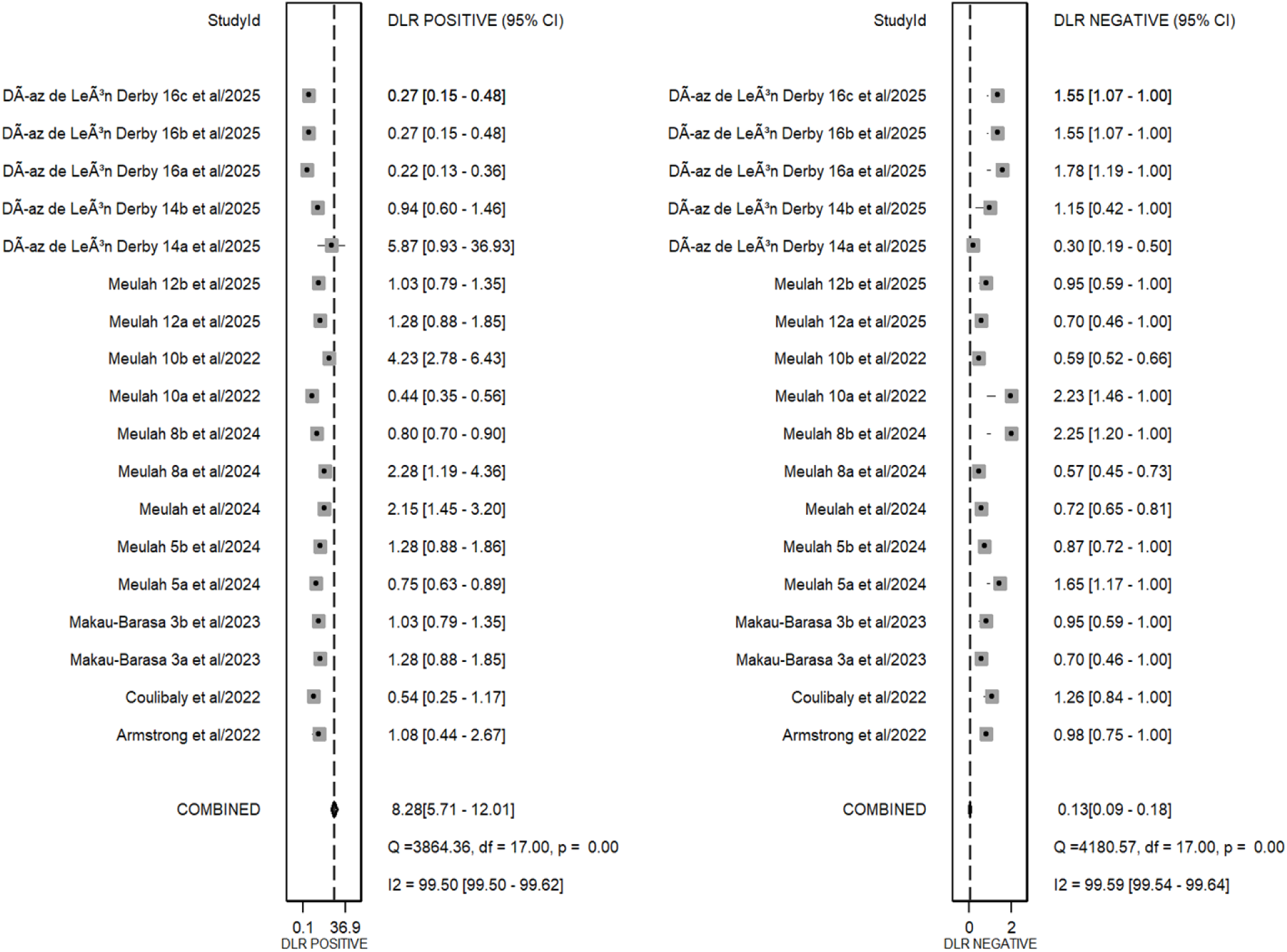
Forest plot illustrates the PLR and NLR of AI-assisted diagnostic tools in the discrimination of positive and negative patients.

**Figure 5:**
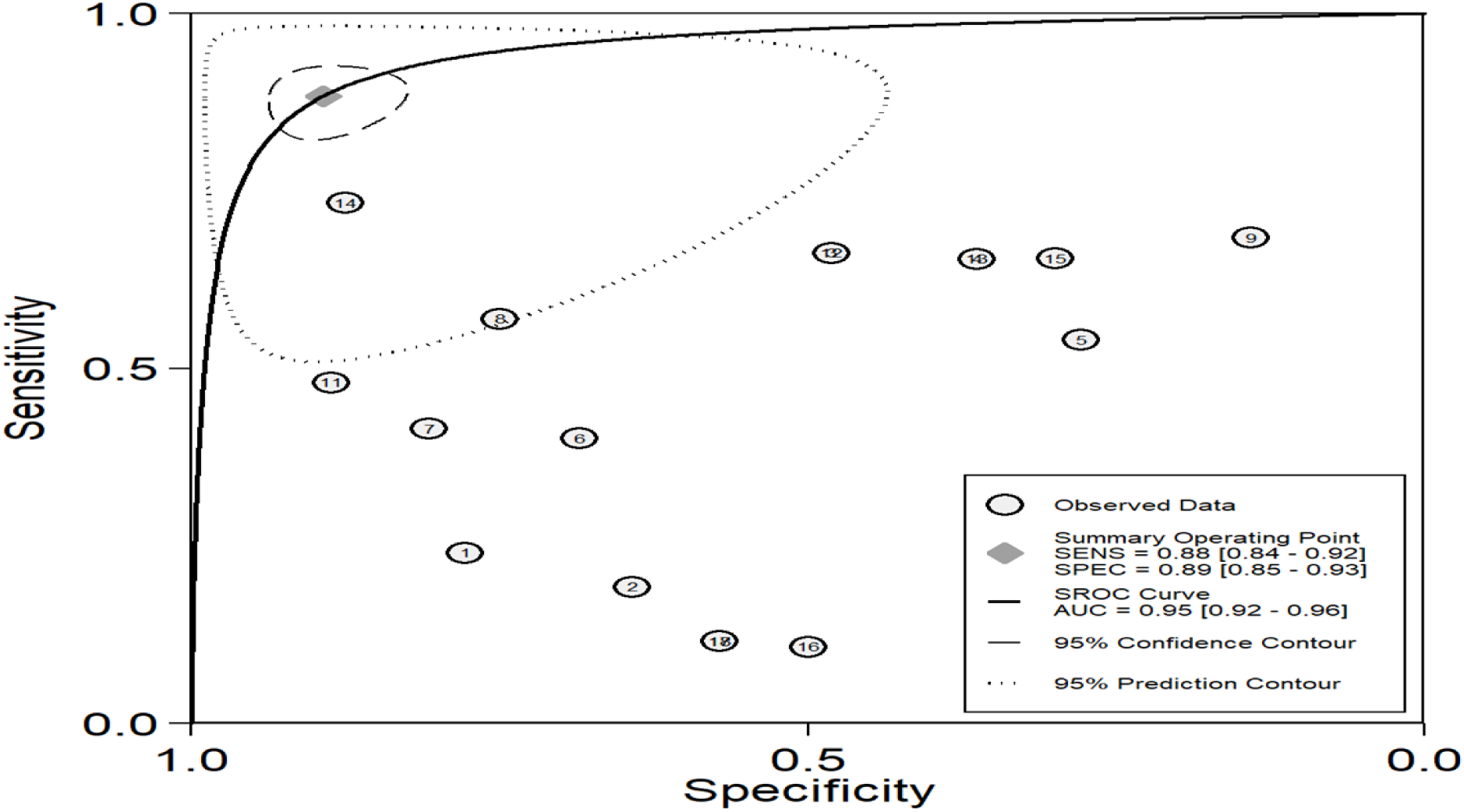
Diagram of SROC curves illustrating the diagnostic performance of AI-assisted diagnostic tools in discriminating between infected and non-infected individuals.

**Figure 6:**
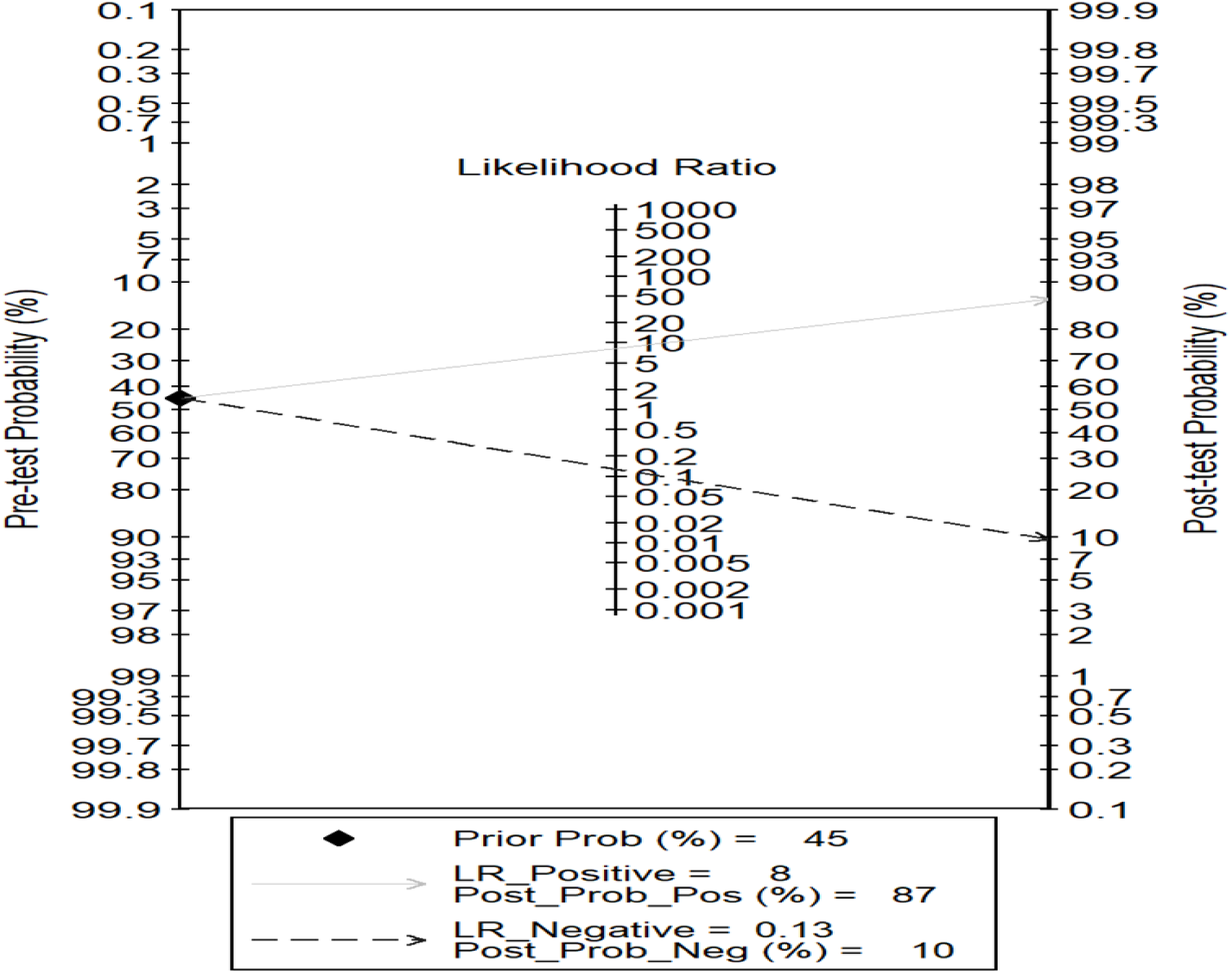
The Fagan nomogram illustrates the capacity of AI-assisted diagnostic tools testing to either confirm or exclude Schistosomiasis in patients.

### Subgroup analysis

#### AI-assisted tools by reference standards

When using urine filtration microscopy as the reference standard (16 studies), the pooled sensitivity and specificity were both 0.9 (95% CI: 0.87–0.93 and 0. 85–0.93, respectively). Despite the high accuracy, heterogeneity was substantial (I² = 99%, p < 0.01), suggesting considerable variability across studies.

When the composite reference standard (CRS) incorporating qPCR and UCP-LF CAA, was used (2 datasets from Gabon), the pooled sensitivity and specificity were 0.89 (95% CI: 0.82-0.94) and 0.88 (95% CI: 0.80-0.93), respectively. This indicates that AI-assisted diagnostic tools maintain high accuracy even when compared against more sensitive molecular and antigen detection methods.

#### By Endemicity Level

In moderate-to-high endemic settings (e.g., Gabon and Nigeria; n = 7 studies), the pooled sensitivity was 0.90 (95% CI: 0.87-0.93) and specificity was 0.88 (95% CI: 0.84-0.92). However, heterogeneity remained high (I² = 97%), likely due to differences in population composition and diagnostic platforms used.

In low endemic settings (e.g., Côte d’Ivoire; n = 3 studies), the pooled sensitivity was 0.86 (95% CI: 0.79-0.91) and specificity was 0.93 (95% CI: 0.88-0.96), with heterogeneity (I² = 88%). These findings suggest that AI-assisted diagnostics retain good specificity even in low-transmission settings, where traditional microscopy often shows reduced sensitivity.

#### By AI-assisted diagnostic tools

Performance varied by AI tool type. SchistoScope-based studies (n = 9) showed a sensitivity of 86% and specificity of 86%, but results were heterogeneous (I² = 98%). AiDx-based studies (n = 4) demonstrated higher accuracy, with pooled sensitivity of 93% and specificity of 91%. This suggests that AiDx may currently outperform SchistoScope in diagnostic applications (Table 2).

**Table 2:** Subgroup analysis of the diagnostic value of AI-assisted diagnostic tools in discriminating against infected and non-infected individuals.

| Subgroup | | No of datasets | Pooled sensitivity (95% CI) | Pooled specificity (95% CI) | Heterogeneity test ($I^2$) | P-value |
| --- | --- | --- | --- | --- | --- | --- |
| Reference test | Urine filtration microscopy | 16 | 90% (87%-93%) | 90% (85%-93%) | 99% | <0.01* |
|  | CRS | 2 | 89% (82%-94%) | 88% (80%-93%) | NA |  |
| Publication year | 2022 | 4 | 85% (80%-88%) | 88% (68%-96%) | 98% | <0.01* |
|  | <2025 | 11 | 88% (82%-92%) | 87% (80%-92%) | 98% | <0.01* |
|  | 2025 | 7 | 90% (82%-94%) | 93% (88%-96%) | 93% | <0.01* |
| Endemicity | Moderate to high endemic setting | 11 | 90% (87%-93%) | 88% (84%-92%) | 97% | <0.01* |
|  | Lower endemic setting | 7 | 86% (79%-91%) | 93% (88%-96%) | 88% | <0.01* |
| AI-assisted diagnostic tools | SchistoScope (AI) | 9 | 86% (79%-91%) | 86% (77%-91%) | 98% | <0.01* |
|  | AiDx | 4 | 93% (92%-95%) | 91% (88%-93%) | 100% | 0.5 |

Note: \* statistically significant, NA-not applicable

#### Meta-regression analysis

The sensitivity of AI-assisted diagnostic tools ranged from 46% to 72%, with the highest values observed in certain AI models and population groups, while specificity ranged from 36% to 50%. Sensitivity differed significantly by population (p = 0.01) and AI model type (p = 0.01), but not by study year or country. Heterogeneity was substantial for study population (I² = 99%) and AI model groups (I² = 98%), but minimal across study years (I² = 0%). These results suggest that diagnostic accuracy is influenced by population characteristics and the AI model employed (Figure 7).

**Figure 7:**
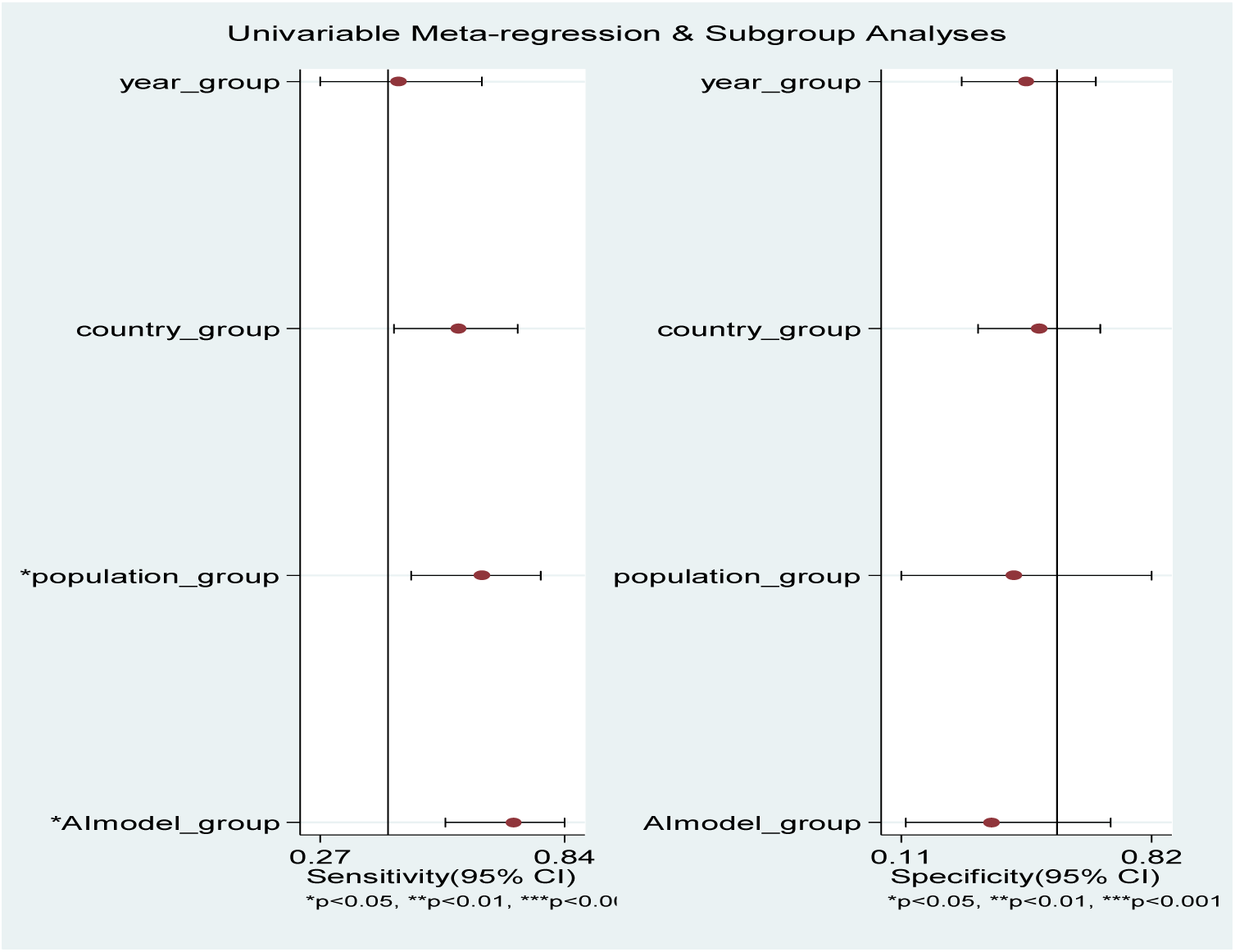
Univariable meta-regression and subgroup analyses of diagnostic accuracy of AI-assisted tools for *Schistosoma haematobium*.

#### Publication Bias

To assess publication bias, Deeks’ funnel plot asymmetry test was conducted. The funnel plot (Figure 8) displayed the relationship between the diagnostic odds ratio (DOR) and study size. The distribution of studies appeared symmetrically around the regression line. The test yielded a *p*-value of 0.65, which is non-significant. This indicates no statistically significant evidence of publication bias among the studies included.

**Figure 8:**
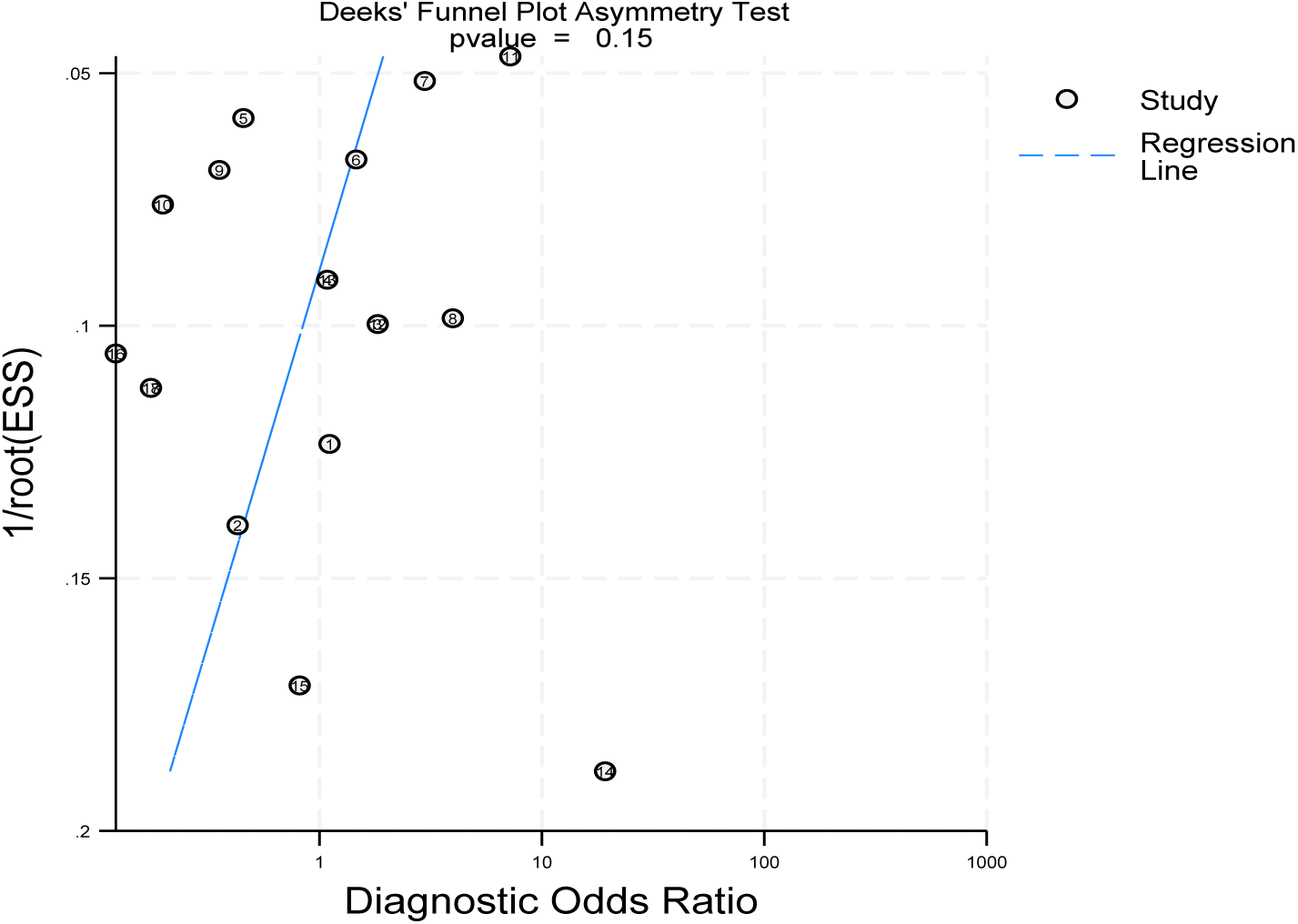
Deek’s funnel plot for publication bias analysis.

## Discussion

The present review demonstrates that AI-assisted microscopy for *S. haematobium* achieves very high diagnostic accuracy. Pooled estimates show a sensitivity of 0.88 and specificity of 0.89. The DOR 64 (95% CI: 38–106) and area under the ROC curve (0.95) both indicate excellent overall discriminative power. In practical terms, the positive likelihood ratio (28.9) and negative likelihood ratio (0.03) imply that a positive AI-assisted test result would greatly increase the post-test probability (from 20% to 67%) while a negative result would reduce it to 3%. Thus, AI tools provide strong rule-in and rule-out capability. However, we note very high between-study heterogeneity (I² = 99%) in pooled estimates.

When compared to traditional tests, AI performance is at least comparable to the best available methods. Urine filtration and light microscopy remain the reference standard, but it is well-recognized that microscopy lacks sensitivity, especially in low-intensity infections [26, 27]. PCR on urine or stool has been shown to be far more sensitive, pooled analyses report 97% sensitivity and 94% specificity for real-time PCR versus microscopy [28]. Our AI pooled sensitivity (88%) approaches that of PCR and exceeds that of conventional microscopy in practice. Likewise, circulating antigen tests offer complementary performance, laboratory-based UCP-LF CAA essays are highly sensitive. In one community survey, 69% of infections were CAA-positive [27].

By comparison, dipstick reagent strips for microhematuria have pooled sensitivity 81% and specifically 89%, but their sensitivity falls (65%) in low-prevalence or post-treatment settings [29]. 88% AI tools sensitivity thus outperforms dipsticks and comes close to PCR sensitivity 97% [28].

In this review, we observed substantial heterogeneity (I²=99%) among studies. This was anticipated given the diversity of AI models, sample preparation, reference standards, and endemic settings. Meta-regression and subgroup analyses suggest variability by population and technology. For instance, community-based surveys tended to show higher sensitivity (93%) than small clinical studies, and the AiDx platform outperformed the Schistoscope in pooled analysis. Differences in slide preparation, microscope quality, and image pre-processing also contributed [28].

Variations in diagnostic performance among AI platforms and populations are mainly due to technological and epidemiological differences. Platform disparities arise from hardware design, image acquisition, and algorithm type. The SchistoScope, which uses smartphone-based imaging, is affected by variations in optical resolution, lighting, and sample alignment, leading to image noise and reduced egg detectability [17]. In contrast, AiDx Assist applies fixed-focus, motorized imaging with uniform illumination and deeper convolutional neural networks (CNNs), producing more consistent and accurate results [18, 22]. Differences in algorithm architecture, such as YOLOv8 versus classical CNNs, also influence feature extraction and noise tolerance, affecting detection rates [20].

Performance variation between community-based and clinical studies reflects differences in infection intensity and sample quality. Community surveys capture a wider infection spectrum, allowing better algorithm training and generalization, while clinical samples often contain fewer eggs or degraded specimens, reducing sensitivity [12, 28]. Community settings typically use fresh urine, whereas clinical samples may be stored or transported, leading to reduced egg visibility [18]. Hence, heterogeneity across studies reflects both genuine biological variation and technical differences. Standardized imaging protocols, open-access training datasets, and external validation are essential to improve reproducibility and scalability of AI-assisted diagnostics.

## Conclusion

AI-assisted image analysis for *Schistosoma haematobium* demonstrated high diagnostic accuracy, with pooled sensitivity and specificity of 0.88 and 0.89, respectively. These findings indicate that AI-based tools can complement existing diagnostic methods for urogenital schistosomiasis.

To ensure reliable and scalable use in endemic settings, further large-scale field validation, standardization of diagnostic protocols, and integration into national control programs are recommended. Future research should focus on algorithm robustness, cost-effectiveness, and usability in routine surveillance, supporting global efforts toward schistosomiasis control and elimination.

## Abbreviations

AI: Artificial Intelligence
AUC: Area Under the curve
CAA: Circulating Anodic Antigen
CNN: Convolutional Neural Network
CRS: Composite Reference Standard
DOR: Diagnostic Odds Ratio
LR+: Positive Likelihood Ratio
LR−: Negative Likelihood Ratio
NTD: Neglected Tropical Diseases
PCR: Polymerase Chain Reaction
PLR: Positive Likelihood Ratio
PIRD: Population-AI-assisted diagnostic tools-Reference Standard-Diagnosis of interest
QUADAS-2: Quality Assessment of Diagnostic Accuracy Studies, version 2
ROC: Receiver Operating Characteristic Curve
SROC: Summary Receiver Operating Characteristic Curve
WHO: World Health Organization
YOLO: You Only Look Once

## Acknowledgements

Not applicable

## Data Availability

Template data collection forms; data extracted from included studies; data used for all analyses; and the review protocol are available from the corresponding author.

## Ethical Approval

Not applicable

## Authors’ contributions

Sisay Desale conceived and designed the study, developed the search strategy, study selection, data extraction and coordinated the review process. He also contributed to data analysis and interpretation and drafted the manuscript.

Getaneh Alemu participated in quality assessment and contributed to the statistical analysis and interpretation of findings. He critically reviewed and revised the manuscript for important intellectual content.

Tadesse Hailu contributed to the development of the methodology, supervised the analysis, provided expert input on interpretation of results, and reviewed the manuscript for scientific accuracy and clarity.

All authors read and approved the final version of the manuscript and agree to be accountable for all aspects of the work.

